# Adverse effects and antibody titers in response to the BNT162b2 mRNA COVID-19 vaccine in a prospective study of healthcare workers

**DOI:** 10.1101/2021.06.25.21259544

**Authors:** Si’Ana A. Coggins, Eric D. Laing, Cara H. Olsen, Emilie Goguet, Matthew Moser, Belinda M. Jackson-Thompson, Emily C. Samuels, Simon D. Pollett, David R. Tribble, Julian Davies, Luca Illinik, Monique Hollis-Perry, Santina E. Maiolatesi, Christopher A. Duplessis, Kathleen F. Ramsey, Anatalio E. Reyes, Yolanda Alcorta, Mimi A. Wong, Gregory Wang, Orlando Ortega, Edward Parmelee, Alyssa R. Lindrose, Andrew L. Snow, Allison M.W. Malloy, Andrew G. Letizia, John H. Powers, Timothy H. Burgess, Christopher C. Broder, Edward Mitre

**Affiliations:** Department of Microbiology and Immunology, Uniformed Services University of the Health Sciences, Bethesda, MD, USA; Henry M Jackson Foundation for the Advancement of Military Medicine, Inc., Bethesda, MD, USA; Department of Preventive Medicine & Biostatistics, Uniformed Services University of the Health Sciences, Bethesda, MD, USA; Infectious Diseases Clinical Research Program, Department of Preventive Medicine and Biostatistics, Uniformed Services University, Bethesda, MD, USA; Clinical Trials Center, Naval Medical Research Center, Silver Spring, MD, USA; General Dynamics Information Technology, Falls Church, VA, USA; Department of Pharmacology and Molecular Therapeutics, Uniformed Services University of the Health Sciences, Bethesda, MD, USA; Department of Pediatrics, Uniformed Services University of the Health Sciences, Bethesda, MD, USA; Infectious Disease Directorate, Naval Medical Research Center, Silver Spring, MD, USA; Clinical Research Directorate, Leidos Biomedical Research, Inc., Frederick National Laboratory for Cancer Research, Frederick, MD, USA

## Abstract

**Background:** mRNA COVID-19 vaccines are playing a key role in controlling the COVID-19 pandemic. The relationship between post-vaccination symptoms and strength of antibody responses is unclear.

**Objective:** To determine whether adverse effects caused by vaccination with the Pfizer/BioNTech BNT162b2 vaccine are associated with the magnitude of vaccine-induced antibody levels.

**Design:** Single center, prospective, observational cohort study.

**Setting:** Participants worked at Walter Reed National Military Medical Center and were seen monthly at the Naval Medical Research Center Clinical Trials Center.

**Participants:** Generally healthy adults that were not severely immunocompromised, had no history of COVID-19, and were seronegative for SARS-CoV-2 spike protein prior to vaccination.

**Measures:** Severity of vaccine-associated symptoms was obtained through participant completed questionnaires. Testing for IgG antibodies against SARS-CoV-2 spike protein and receptor binding domain was conducted using microsphere-based multiplex immunoassays.

**Results:** 206 participants were evaluated (69.4% female, median age 41.5 years old). We found no correlation between vaccine-associated symptom severity scores and vaccine-induced antibody titers one month after vaccination. We also observed that 1) post-vaccination symptoms were inversely correlated with age and weight and more common in women, 2) systemic symptoms were more frequent after the second vaccination, 3) high symptom scores after first vaccination were predictive of high symptom scores after second vaccination, and 4) older age was associated with lower titers.

**Limitations:** Study only observes antibody responses and consists of healthy participants.

**Conclusions:** Lack of post-vaccination symptoms following receipt of the BNT162b2 vaccine does not equate to lack of vaccine-induced antibodies one month after vaccination. This study also suggests that it may be possible to design future mRNA vaccines that confer robust antibody responses with lower frequencies of vaccine-associated symptoms.

**Funding:** This study was executed by the Infectious Disease Clinical Research Program (IDCRP), a Department of Defense (DoD) program executed by the Uniformed Services University of the Health Sciences (USUHS) through a cooperative agreement by the Henry M. Jackson Foundation for the Advancement of Military Medicine, Inc. (HJF). This project has been funded by the Defense Health Program, U.S. DoD, under award HU00012120067. Project funding for JHP was in whole or in part with federal funds from the National Cancer Institute, National Institutes of Health, under Contract No. HHSN261200800001E. The funding bodies have had no role in the study design or the decision to submit the manuscript for publication.

## Introduction

The recent implementation of mRNA-based SARS-CoV-2 vaccines is playing a major role in efforts to control the SARS-CoV-2 pandemic. Both the Pfizer/BioNTech BNT162b2 and Moderna mRNA-1273 induce high titer anti-SARS-CoV-2 antibodies and confer robust protection against morbidity and mortality from SARS-CoV-2 infection (1-4).

One feature of the SARS-CoV-2 mRNA vaccines is their high level of reactogenicity, with both local and systemic reactions reported by the majority of recipients in Phase 1-3 studies (1-4). A Centers for Disease Control and Prevention vaccine safety monitoring program of adverse effects (AEs) in the U.S. population has found that injection site pain (79.3%), fatigue (53.5%), myalgia (47.2%), headache (43.4%), chills (30.6%), fever (29.2%), and joint pains (23.5%) are frequent after the 2^nd^ dose of the BNT162b2 vaccine (5).

Reactogenicity to vaccines is typically driven by activation of the innate immune system through ligation of pattern-recognition receptors and subsequent release of inflammatory cytokines such as interleukin-1, interleukin-6, and tumor necrosis factor (6). Studies suggest type I interferon production elicited by direct mRNA recognition is critical for SARS-CoV-2 control (7-10), and likely contributes to both immunogenicity and reactogenicity of SARS-CoV-2 mRNA vaccines (6). Adaptive immune pathways also likely play a role in causing vaccine-mediated symptoms, especially during booster vaccinations or vaccination following infection.

During the roll-out of COVID-19 vaccines, it has become commonplace for media outlets and medical professionals to state that presence of symptoms means that a vaccine is “working” (11, 12). Although this statement is fundamentally true because vaccines “work” by inducing inflammatory responses, it also implies incorrectly that lack of symptoms post-vaccination may indicate an absence of appropriate antiviral antibody responses. Notably, there is little data demonstrating correlations between vaccine-induced symptoms and antibody titers with any vaccine platforms (6, 13). The goal of this study was to assess for correlation between AEs caused by BNT162b2 vaccination and the magnitude of SARS-CoV-2 antibody responses one month after second vaccination dose.

## Methods

### Study participants

Participants were enrolled in the Prospective Assessment of SARS-CoV-2 Study (PASS), an observational, longitudinal cohort study of healthcare workers (HCWs) that is evaluating clinical and immunological responses to SARS-CoV-2 infection and vaccination. The cohort consists of generally healthy adults who are ≥ 18 years old, work at Walter Reed National Military Medical Center (WRNMMC), are not severely immunocompromised, and were seronegative for SARS-CoV-2 at time of study enrollment. Details of inclusion and exclusion criteria can be found in the protocol, which has been published (14). The subset of PASS participants included for analysis in this study also met the following criteria: (i) no history of COVID-19 diagnosis, (ii) seronegative for SARS-CoV-2 anti-spike protein IgG prior to vaccination, (iii) received two vaccinations with the Pfizer/BioNTech BNT162b2 vaccine, and (iv) completed two vaccination symptom questionnaires by March 24, 2021. PASS was initiated in August of 2020 with study participants seen monthly at the Naval Medical Research Center Clinical Trials Center. The study protocol was approved by the Uniformed Services University Institutional Review Board.

### Assessment of vaccine-associated symptoms

Participants completed a structured vaccine-associated symptoms questionnaire at the first monthly visit after each vaccination dose. Questionnaires asked about the presence and severity of 12 symptoms (8 categorized as systemic, 3 categorized as localized to the vaccine site, and 1 categorized as non-local and non-systemic, see Table 1). Severity of each symptom was defined as symptom intensity and measured on a scale of 0 – 4 (0 = not at all, 1 = a little bit, 2 = somewhat, 3 = quite a bit, 4 = a lot), with scores for each symptom summed for a total symptom severity score of 0 – 48. Participants were also asked to report the total duration of any vaccine-associated symptoms.

**Table 1:** Demographic characteristics of participants in the full cohort and in the titers subgroup.

| Characteristic | Full Cohort<br>N=206 | Titers Subgroup<br>n=101 |
| --- | --- | --- |
| **Sex—no. (%)** |  |  |
| Female | 143 (69.4) | 68 (67.3) |
| Male | 63 (30.6) | 33 (32.7) |
| **Ethnicity—no. (%) †** |  |  |
| Hispanic | 10 (4.9) | 1 (1.0) |
| Not Hispanic | 193 (93.7) | 100 (99.0) |
| not reported | 3 (1.5) | 0 (0) |
| **Race—no. (%) †** |  |  |
| White | 150 (72.8) | 79 (78.2) |
| Black | 23 (11.2) | 10 (9.9) |
| Asian | 21 (10.2) | 10 (9.9) |
| Native Hawaiian/Other Pacific Islander | 1 (0.5) | 0 (0) |
| Multiracial | 6 (2.9) | 2 (2.0) |
| Not reported | 5 (2.4) | 0 (0) |
| **Age—yr ‡** |  |  |
| Mean (IQR) | 42.4 (33.0-51.3) | 43.2 (34.0-52.0) |
| Median (Range) | 41.5 (20-69) | 43.0 (20-69) |
| **Weight—kg** |  |  |
| Mean (IQR) | 76.1 (63.7-86.4) | 76.1 (63.5-84.3) |
| Median (Range) | 73.0 (42.5-141.2) | 71.3 (42.5-135.7) |
| Female Mean (IQR) | 70.4 (60.5-76.5) | 68.0 (60.1-74.2) |
| Male Mean (IQR) | 89.2 (76.2-97.0) | 92.8 (78.9-111.4) |

† Ethnic and racial group was reported by the participants
‡ The age of the participant was the age at the time of PASS study enrollment

### Antibody testing

IgG antibodies against SARS-CoV-2 spike protein and receptor binding domain (RBD) were measured using microsphere-based multiplex immunoassays (MMIAs) built using Luminex xMAP-based technology as previously described (15) (Supplemental Methods).

### Statistical analyses

Wilcoxon signed rank test was used for paired comparisons and Mann-Whitney for unpaired comparisons. Kruskall-Wallis or ANOVA was used when comparing multiple groups. Spearman rank analyses were used to assess for correlations. Spearman partial correlations were used to determine if age, sex, or weight were independently associated with vaccine-associated symptom scores and to adjust for age, sex, and weight when assessing for correlations between vaccine-related symptom scores and antibody titers.

## Results

### Study participants evaluated

A total of 206 participants, of 271 enrolled in the PASS study, were seronegative and without a history of COVID-19 diagnosis when they received the first of two BNT162b2 vaccinations, and provided a serum sample at least three weeks after final vaccination. Of these, the median age was 41.5 years old (IQR 33-51.25) and 69.4% were female. Anti-spike antibody levels were quantified by MMIA mean fluorescence intensity (MFI) for all participants and by endpoint dilution titers for the first 101 participants for which serum at least three weeks after second vaccination dose was available. Demographic information for the study cohort and titers subgroup is in Table 1.

### Symptom severity scores after first and second vaccinations

The mean symptom score reported for the second vaccination was significantly greater than that of the first (10.62, IQR 3-16, vs 7.3, IQR 3-10, p < 0.0001, Figure 1A), even though there was no significant difference in the duration of symptoms following either vaccination (Figure 1B). To better understand the observed difference in symptom severity, participant symptom scores were subdivided into systemic (maximum score: 32) and local (maximum score: 12) symptom scores. Interestingly, while the mean systemic symptom score after the second vaccination was significantly greater than after the first (7.0, IQR 0-12, vs 3.1, IQR 0-5, p < 0.0001) (Figure 1C), local symptoms displayed an opposing trend with lower severity following the second vaccination (mean 3.3, IQR 1-5, vs 4.2, IQR 2-6, p < 0.0001) (Figure 1D). Overall, there was a positive correlation between vaccination 1 and vaccination 2 symptom scores (rho = 0.28, p< 0.0001) (Figure 1E). To determine how frequently individuals with substantial symptoms after the first vaccination develop substantial symptoms after the second vaccination, participants were separated into two groups: a low symptom severity group made up of individuals with vaccination 1 symptom scores less than or equal to 10 (n=164, 79.6%) and a high severity group comprised of participants with vaccination 1 symptom scores greater than 10 (n=42, 20.4%). Roughly 35% of participants in the low symptom severity group reported a symptom score greater than 10 following the second vaccination (Figure 1F). This frequency nearly doubled in the high severity group, with 64% of those participants having recorded a symptom score greater than 10 following the second vaccination (Figure 1F).

**Figure 1:**
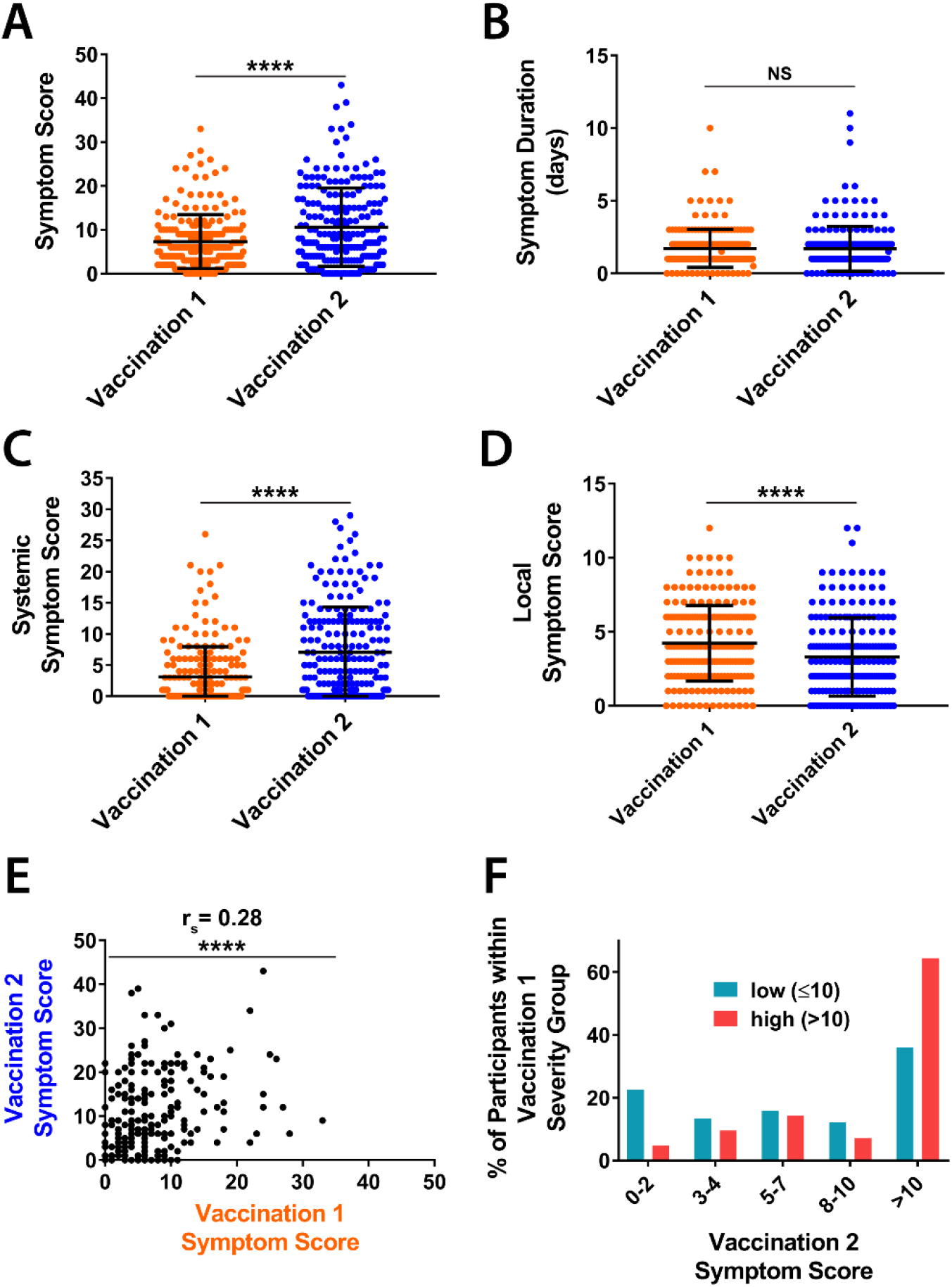
Symptom severity after the first vaccination with BNT162b2 correlates with severity of symptoms following the second vaccination. (A) Total symptom severity scores (range 0 – 48), (B) duration of symptoms, (C) systemic symptom severity scores (range 0-32), and (D) local symptom severity scores (range 0-12) following the first (orange) and second (blue) vaccinations. (E) Correlation of vaccination 1 and vaccination 2 symptom scores. (F) Percentage of subjects categorized as having low (≤10, teal) or high (>10, red) total symptom scores after vaccination 1 that exhibited symptom scores of 0-2, 3-4, 5-7, 8-10, or > 10 after vaccination 2. (N = 206 for all panels, **** = p < 0.0001, NS = not significant, significance assessed by Wilcoxon signed rank test for panels A-D and by Spearman correlation analysis for panel E. Bars represent mean and standard deviation in panels A-D).

### Frequency of specific symptoms experienced after first and second vaccine doses

Soreness at the injection site (vaccination 1: 91.3%, vaccination 2: 82.0%), pain at the injection site (vaccination 1: 71.8%, vaccination 2: 62.1%), and the feeling of being weak or tired (vaccination 1: 42.2%, vaccination 2: 62.1%) were the three most common symptoms reported after receiving the first (Table 2A) and second (Table 2B) doses of the Pfizer-BioNTech BNT162b2 vaccine. Except for the local symptoms of soreness, pain, or redness at the injection site, all symptoms listed on the questionnaire increased in frequency from the first vaccination to the second. For example, while 28.2% of participants experienced body aches or pains following the first vaccination, 52.4% reported this symptom following the second vaccination. This increase was present for even the least common symptom of swollen lymph nodes, which increased from 4.4% to 13.6% of participants following the first and second doses, respectively.

**Table 2A:**
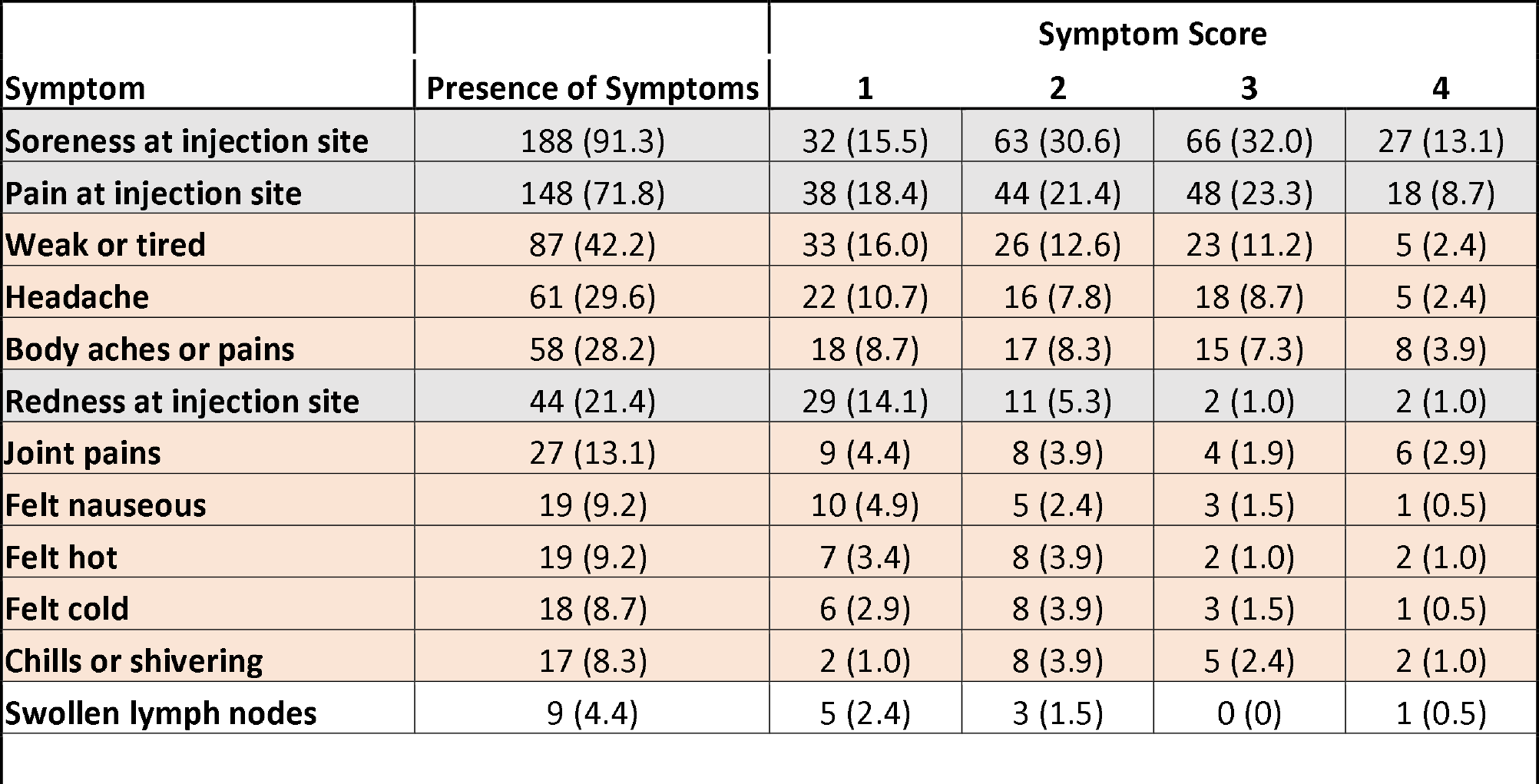
Symptoms experienced after first vaccination, ranked by frequency.

**Table 2B:**
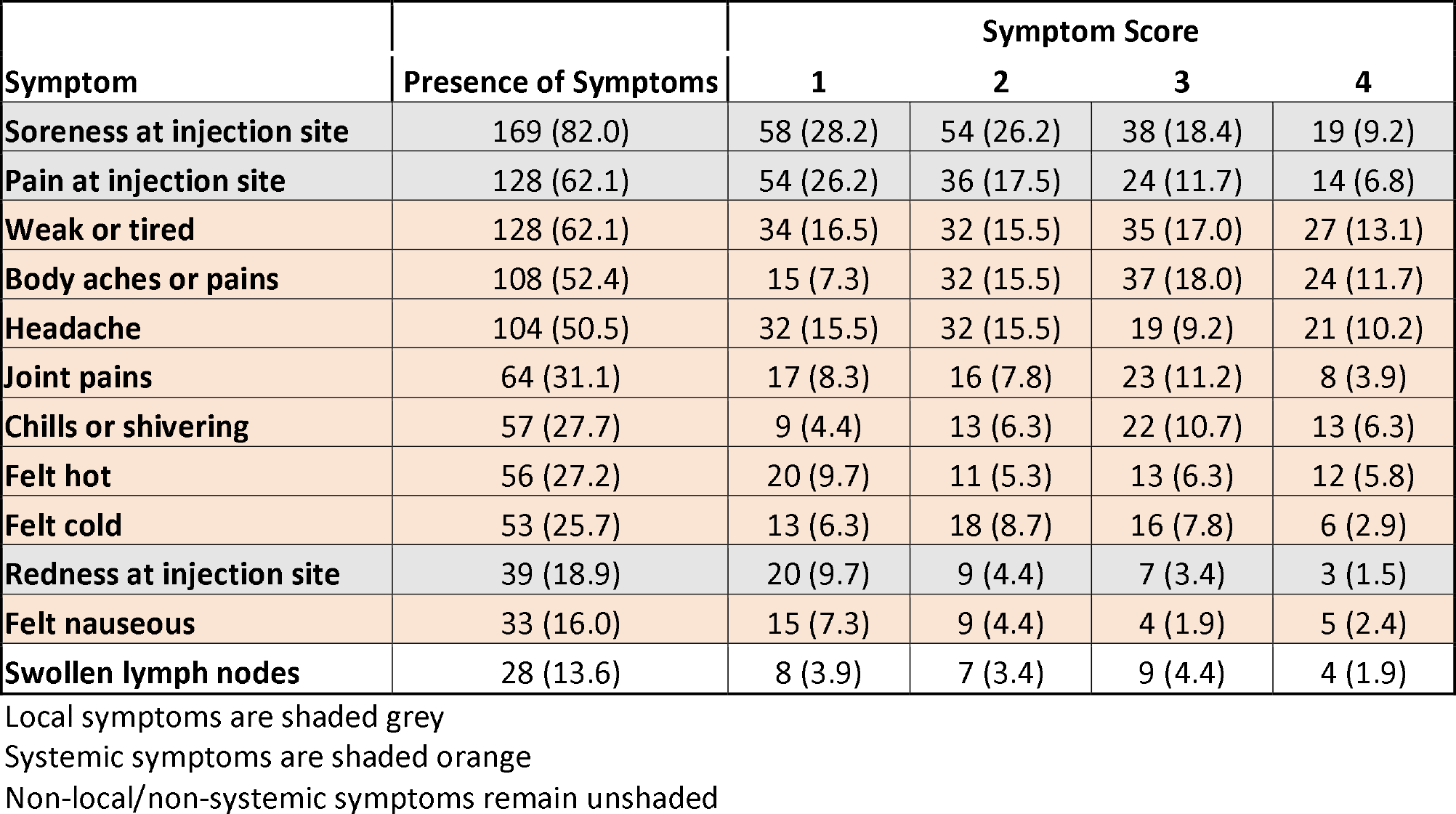
Symptoms experienced after second vaccination, ranked by frequency.

### Relationship between vaccine symptoms, age, sex, and weight

Younger age, female sex, and lower weight were all associated with higher symptom scores when evaluated individually. There was a modest, yet significant, negative correlation between age and symptom severity for both the first (Figure 2A: rho = -0.17, p= 0.02) and second (Figure 2B: rho = -0.17, p= 0.01) vaccine doses. Female participants reported significantly higher symptom scores than males following the first vaccination (mean 8.0, IQR 4-10, vs mean 5.7, IQR 2-9, p = 0.006, Figure 2C). Females also had higher symptom scores after the second vaccination compared to males (mean 11.3, IQR 4-18, vs mean 9.1, IQR 3-15), but this difference was not statistically significant (p=0.11, Figure 2D). Spearman partial correlation analysis determined age to be an independent predictor of total symptom scores after both 1st (partial rho = -0.17, p=0.018) and 2nd (partial rho = 0.17, p=0.018) vaccinations after adjusting for sex and weight (Supplemental Table 1). While not statistically significant, female sex was consistently found to positively correlate with symptom scores and weight to negatively correlate with symptom scores when analyzed with partial Spearman correlations (Supplemental Table 2). No differences in symptom scores based on race for either first or second vaccination were noted (Supplemental Figure 1).

**Figure 2:**
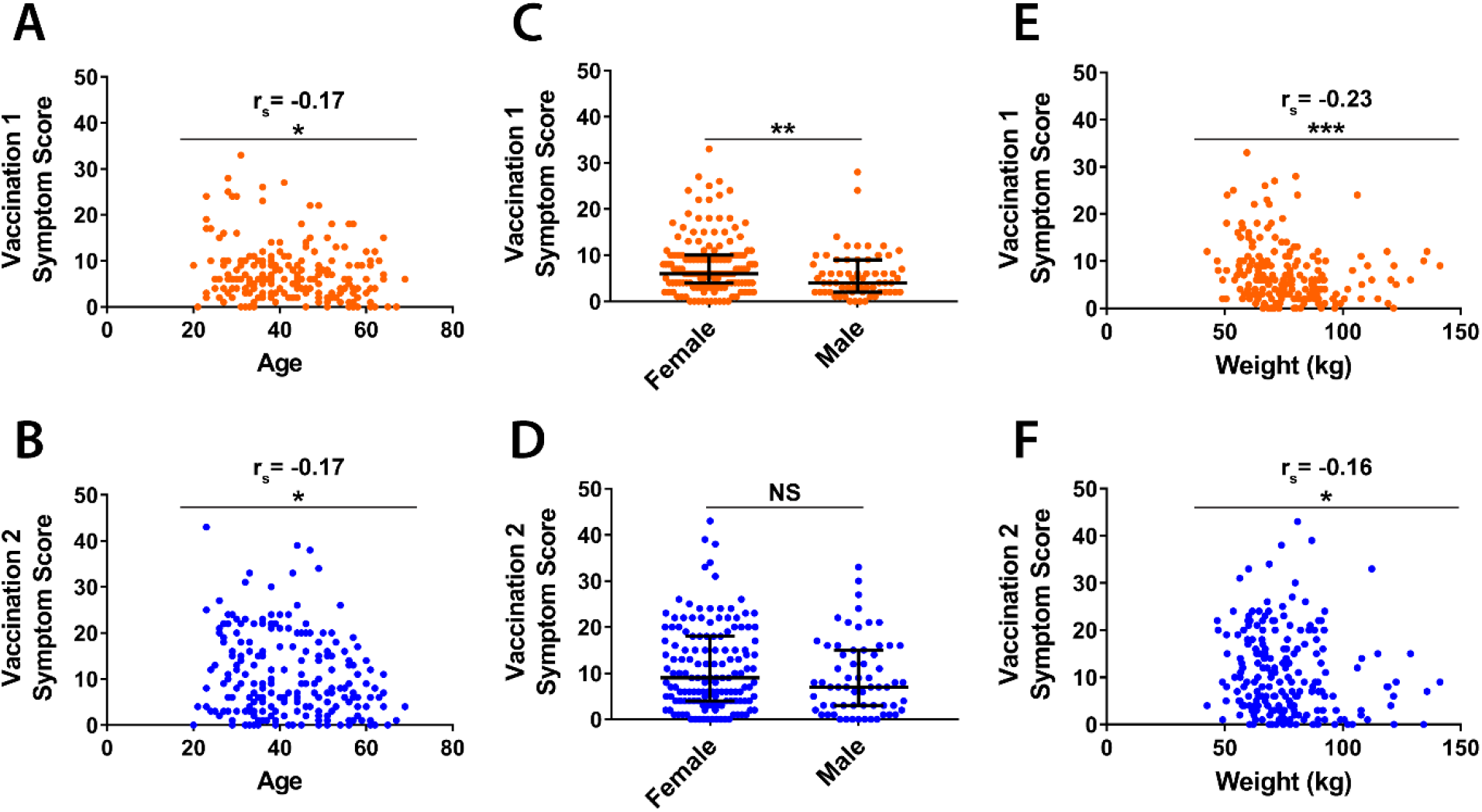
Younger age, female sex, and lower weight are associated with higher symptom severity. Total symptom severity scores after vaccination 1 (orange) and vaccination 2 (blue) graphed against age (A, B), sex (C,D), and weight (E,F). Correlations were assessed by Spearman rank analysis for age and weight. Mann Whitney analysis was used to assess for significance between males and females. (N = 206 for all panels. * = p < 0.05, ** = p < 0.01, *** = p < 0.001, NS = not significant. Bars represent median and IQR in panels C and D).

### Lack of correlation between vaccine-associated symptoms and antibody titers

Time between final vaccination and serum sampling was a mean of 36.8 days (range 22-104, IQR 29-43). Older age, but not sex, weight, or race, was negatively associated with vaccine-induced antibody levels (Figure 3 and Supplemental Figure 2). No correlation between symptom severity following the first or second vaccine doses and IgG reactivity with spike protein was noted (Figure 4 A, B). Endpoint dilution assays also exhibited no correlation between vaccine symptom scores and endpoint titers of anti-spike IgG (Figure 4 C, D) or anti-RBD IgG (Figure 4E, F). Lack of correlation was observed with both Spearman rank analyses and with partial Spearman correlations analyses after adjusting for age and sex. Secondary analyses also revealed no associations between systemic symptoms or lymph node swelling and anti-spike and anti-RBD titers (data not shown). Analysis of total symptom duration after first or second vaccination revealed no association with anti-spike MFI levels or anti-RBD IgG titers, though a significant correlation was observed with duration of symptoms after second vaccination and anti-spike IgG titers (Supplemental Figure 3).

**Figure 3:**
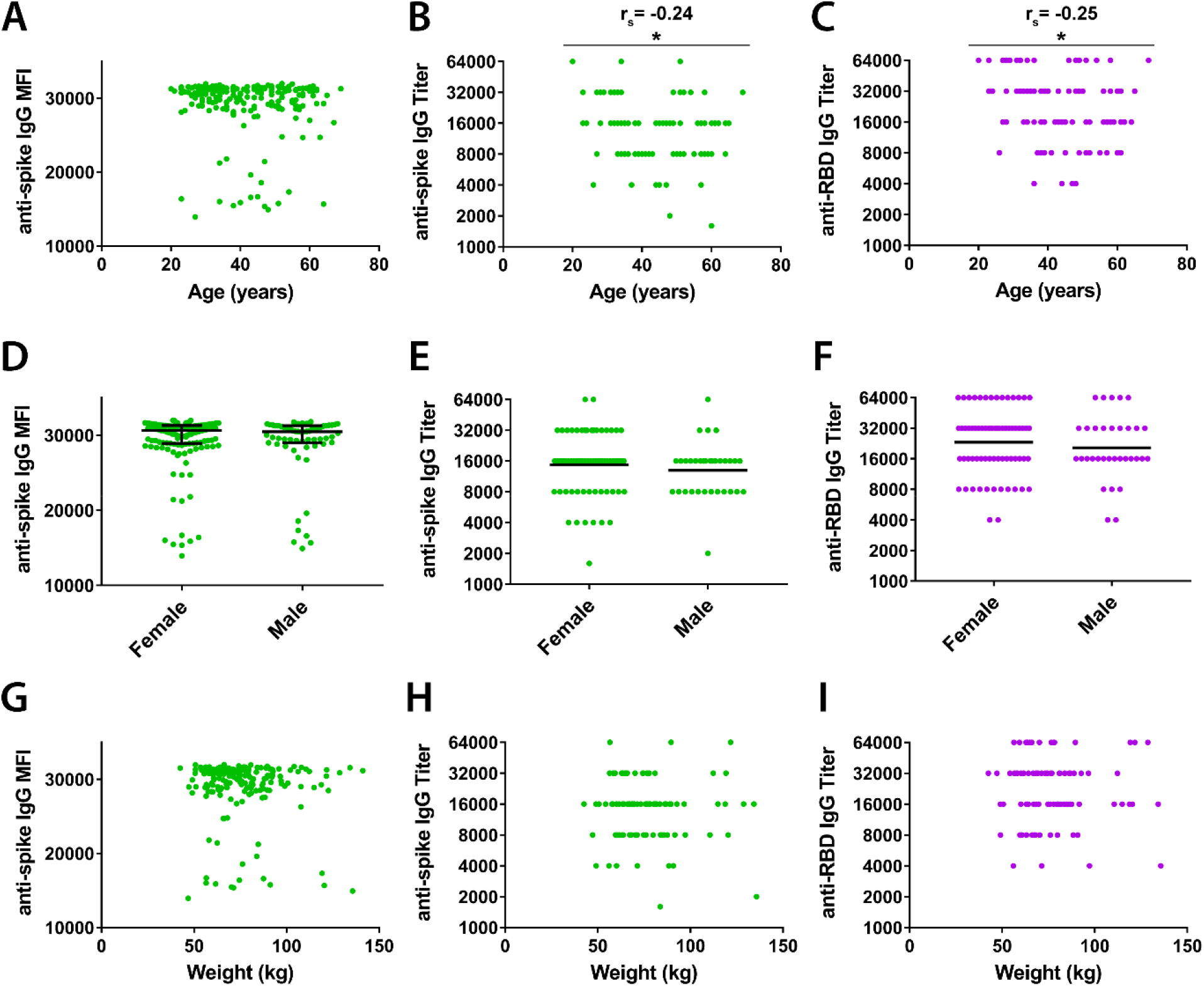
Age, but neither sex nor weight, correlates with BNT162b2-induced anti-SARS-CoV-2 antibody titers. Samples collected one month (mean 36.8 days) after participants received the second vaccine dose were tested for antibodies against SARS-CoV-2 spike protein (green) and receptor binding domain (RBD, purple) and plotted against age (A-C), sex (D-F), and weight (G-I). Antibody reactivity against SARS-CoV-2 spike protein in samples diluted 1:400 was assessed in 206 subjects and reported as mean fluorescence intensity (MFI). Endpoint dilution titers were measured in a subset of 101 subjects for both anti-spike IgG and anti-RBD IgG. Correlations were assessed by Spearman rank analysis for age and weight, and Mann Whitney analysis was used to assess for significance between males and females. (N = 206 for MFI values and n=101 for antibody titers. * = p < 0.05. Bars indicate median and IQR (D) or geometric mean (E, F). Titers recorded as >32000 are plotted as 64000).

**Figure 4:**
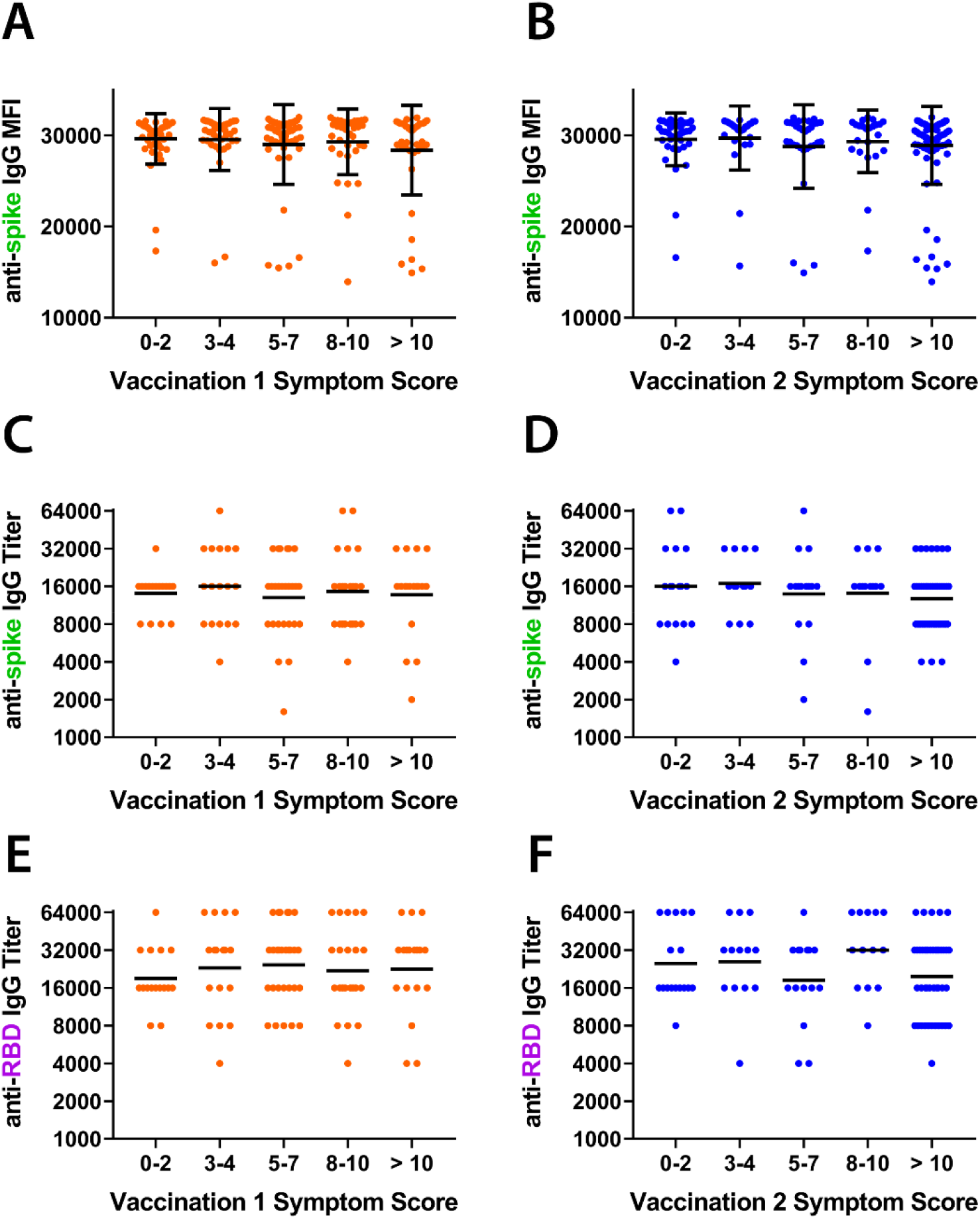
Severity of symptoms after vaccination correlates with neither vaccine-induced anti-Spike IgG reactivity nor with titers of anti-Spike and anti-RBD IgG antibodies. Samples collected one month (mean 36.8 days) after participants received the second vaccine dose of BNT162b2 were tested for antibodies against SARS-CoV-2 spike protein and RBD and plotted against total symptom scores. (A, B) Levels of anti-spike IgG antibodies as measured by MFI after first (orange) and second (blue) vaccination (N=206). (C, D) Titers of anti-spike IgG antibodies after first and second vaccination (n=101). (E, F) Titers of anti-RBD IgG antibodies after first and second vaccination. Bars indicate mean and standard deviation (A,B) or geometric mean (C-F). Titers recorded as >32000 are plotted as 64000. Assessments for correlations were conducted by both Kruskal-Wallis analysis with subjects binned into categories of symptom score ranges and by Spearman rank analysis evaluating antibody levels against symptom scores as a continuous variable. All showed no significant correlations.

## Discussion

Local and systemic symptoms often occur after vaccination and are predominantly due to activation of inflammatory pathways (6). In this study, we evaluated vaccine-related AEs that occur in response to the Pfizer/BioNTech BNT162b2 mRNA COVID-19 vaccine in a cohort of healthcare workers. We observed that 1) symptoms were more common in women and inversely correlated with age and weight, 2) systemic symptoms were more frequent after the 2^nd^ vaccination, 3) high symptom scores after first vaccination were associated with high symptom scores after second vaccination, and 4) older age was associated with lower vaccine-induced antibody titers. Notably, we found no correlation between vaccine-related symptom severity and vaccine-induced antibody titers. This lack of correlation was observed even when adjusting for age, weight, and sex.

Local symptoms were more frequent than systemic symptoms after both first and second vaccinations, with pain, soreness, and redness reported in 91%, 72%, and 21% of participants, respectively, after first vaccination and by 82%, 62%, and 19% of participants, respectively, after the second vaccination. These frequencies paralleled those observed in the clinical trials of BNT162b2 (3, 4). Systemic symptoms were common with symptoms of feeling weak or tired, having body aches or pains, or having joint pains reported by 42%, 28%, and 13% of participants, respectively, after first vaccination and by 62%, 52%, and 31%, respectively, after the second vaccination. Again, these frequencies were similar to clinical trial reports (3).

Partial correlation analysis demonstrated that age was an independent predictor of vaccine-related AEs, with age exhibiting a rho factor of – 0.17 for total symptom scores after both vaccine doses in bivariate analyses after controlling for sex and weight. The finding that older age is associated with lower post-vaccination symptoms is similar to results of BNT162b2 clinical trials and to findings reported in a comparison study evaluating vaccine responses in individuals greater than 80 years old and less than 60 years old (3, 4, 16).

While both women and individuals with lower weights were found to have greater vaccine-related symptoms in our study, neither sex nor weight was found to be an independent predictor of symptom scores. We speculate that our study may have been insufficiently powered to separate the independent contributions of sex and weight to vaccine-associated symptoms, although sex and weight may simply not be independent predictors. Consistent with our findings, women in a large-scale United Kingdom study were found to have more symptoms than men after BNT162b2 vaccination (17). Additionally, women have been shown to have greater reactogenicity to other vaccines, including measles/mumps/rubella, hepatitis B, influenza, and yellow fever (18, 19). The determination of whether women and/or low weight individuals have greater BNT162b2 AEs may be informed by additional cohort studies.

The mechanisms by which younger individuals, or women, exhibit greater vaccine-related AEs is unclear, but may be due to differences in innate immune function. Dendritic cells of older individuals have been shown to release decreased quantities of pro-inflammatory cytokines when stimulated through pattern recognition receptors (20, 21). For women, increased AEs may be due to increased responsiveness of innate immune pathways, though differences in anatomy at injection sites, sex hormones, and adaptive immune function may also play a role (22, 23).

Of note, in addition to being associated with lower vaccine-associated symptom scores, age was also found to be significantly correlated with lower titers of vaccine-induced IgG antibodies against spike protein and RBD. Reduced titers were also observed in elderly individuals in both the phase I clinical trial of BNT162b2 and in a recent study evaluating BNT162b2 responses in individuals greater than 80 years old (3, 16). The mechanisms underlying reduced antibody responses in elderly individuals are not yet fully elucidated, but likely include factors such as reductions in T cell receptor signaling, predilection for naïve T-cells to differentiate into effector rather than memory T cells, decreased function of follicular helper T cells, and lower antibody production by plasma cells (24).

In regards to vaccination reactogenicity, as with the BNT162b2 clinical trials (3, 4), we observed greater symptom severity after the second vaccination. Notably, individuals that had a high symptom score after the first vaccination were almost twice as likely to have substantial symptoms after second vaccination compared to those with a low symptom score from the first vaccination. Nevertheless, individuals with few symptoms after the first vaccination still had a 35% chance of having substantial symptoms (total symptom score > 10) after second vaccination.

Finally, we did not observe a significant correlation between vaccine-related AEs and the magnitude of vaccine-induced antibody titers. Individuals with both high and low symptom scores had similar levels of spike-specific IgG antibodies when measured by MFI, and similar endpoint dilution titers of both spike-specific and RBD-specific IgG antibodies. This lack of correlation was maintained even when controlling for age and sex. While it would appear logical that vaccine-associated AEs could be predictive of antibody titers, there is little evidence for such a relationship (6). One study that evaluated different adjuvants for hepatitis B vaccination found a modest association of symptoms after first vaccination with CD4+ T-cell responses (25). However, the same study found no association between first vaccination AEs and antibody responses and no associations between symptoms after second vaccination and either CD4+ T-cell or antibody responses (25). Consistent with our study, Muller et al did not find an association between symptoms induced by BNT162b2 vaccination and antibody titers (16).

The lack of correlation between vaccine-associated symptoms and antibody titers has two important implications for mRNA SARS-CoV-2 vaccines. First, individuals that exhibit few symptoms after vaccination can be reassured that this does not mean the vaccine “didn’t work.” Indeed, in this cohort individuals with few to no symptoms were just as likely to have developed strong antibody responses as individuals that exhibited substantial symptoms. Second, the immunological pathways responsible for mRNA vaccine-induced AEs may not be required for development of robust antibody responses.

mRNA vaccines induce inflammation through multiple pathways, including ligation of innate immune receptors, release of inflammatory cytokines and chemokines, and activation of antigen presenting cells, natural killer cells, and antigen-specific T and B cells (26-31). The exact pathways each mRNA vaccine induces, however, likely varies depending on factors such as the constituents of the lipid nanoparticle, the cellular uptake pathways for the lipid nanoparticle, the amount of contaminating double-stranded RNA, nucleotide modifications, and the antigen being encoded by the delivered mRNA. Given the lack of association between symptoms and antibody titers, we speculate that some pathways may be expendable for development of robust adaptive immune responses. If such pathways can be defined, then efforts on developing mRNA vaccines that minimally activate such pathways could be advantageous.

The present study is limited to antibody responses. It is possible that vaccine reactogenicity, while not impacting antibody response magnitude, might correlate with vaccine-induced antigen-specific T-cell responses. Additionally, whether reactogenicity impacts durability of vaccine-induced immune responses will be an important area to explore going forward. The assessment of both vaccine-induced T-cell and antibody responses over time in the cohort described here is ongoing. Notably, we did find that symptom duration after second vaccination correlated with vaccine-induced anti-spike IgG titers, but not with anti-spike MFI levels or anti-spike RBD titers. This may reflect a true association, or an artifact due to conducting multiple secondary analyses. Another limitation is that the cohort consisted of healthy volunteers without substantial immunocompromising conditions. We anticipate that studies in other cohorts will further inform the relevance of vaccine-associated symptoms to other populations and whether duration of vaccine symptoms correlates with vaccine-induced antibody titers.

In conclusion, this study demonstrates that BNT162b2 vaccinations are commonly associated with both local and systemic symptoms. Symptoms are greater after second vaccination, are more common in younger individuals, and do not correlate with vaccine-induced antiviral IgG titers. These findings suggest that patients receiving the BNT162b2 vaccine should be reassured that lack of symptoms does not necessarily equate to lack of desired vaccine function. This study also suggests that it may be possible to design future mRNA vaccines that confer robust antibody responses with lower frequencies of vaccine-associated symptoms. Indeed, emerging studies suggest the balance between vaccine immunogenicity and reactogenicity can be better tuned for COVID-19 mRNA-based vaccines as well (32).

## Data Availability

Data are available from the first (S.C.) and corresponding (E.M.) authors upon request.

## Acknowledgments

We thank Camille Estupigan, Dr. Mark Simons, and Dr. Kimberly Edgel for providing insightful comments on the manuscript.

