## Supplemental Materials for "Adverse effects and antibody titers in response to the BNT162b2 mRNA COVID-19 vaccine in a prospective study of healthcare workers"

**Supplemental Tables and Figures**

|  | **Total symptom score vaccine 1** | **Systemic symptoms score vaccine 1** | **Local symptoms score vaccine 1** | **Total symptom score vaccine 2** | **Systemic symptoms score vaccine 2** | **Local symptoms score vaccine 2** |
| --- | --- | --- | --- | --- | --- | --- |
| **Female** | .104  (.140) | .058  (.408) | .134  (.056) | .050  (.476) | .049  (.491) | .019  (.788) |
| **Age (per year)** | -.166*  (.018) | -.128  (.068) | -.100  (.155) | -.165*  (.018) | -.126  (.073) | -.107  (.128) |
| **Weight (per kg)** | -.133  (.057) | -.045  (.525) | -.100  (.155) | -.100  (.155) | -.076  (.279) | -.126  (.072) |

**Supplemental Table 1:** Partial Spearman correlation coefficients for female sex, weight, and age with total symptom scores (range 0-48), systemic symptom scores (range 0-32), and local symptom scores (range 0-12) after first and second vaccinations. Coefficients in each row are adjusted for the variables listed in the other rows. For female sex, a positive coefficient indicates that women have higher symptom scores than men. (N = 206. P values are in parentheses. * p < 0.05. ** p < 0.01)

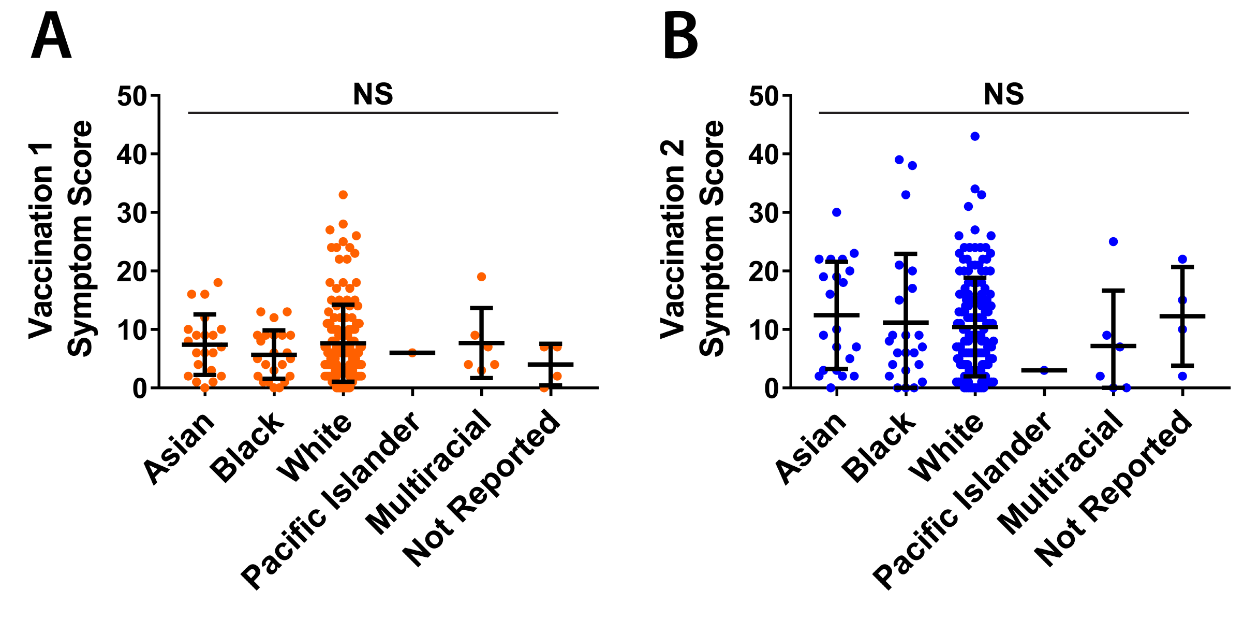

**Supplemental Figure 1: Lack of correlation between race and vaccine-associated symptoms.** Reported symptom scores following the first (A) and second (B) vaccinations plotted by race. (N = 206 subjects. Statistical significance evaluated by ANOVA. NS= no significance while bars represent mean and standard deviation).

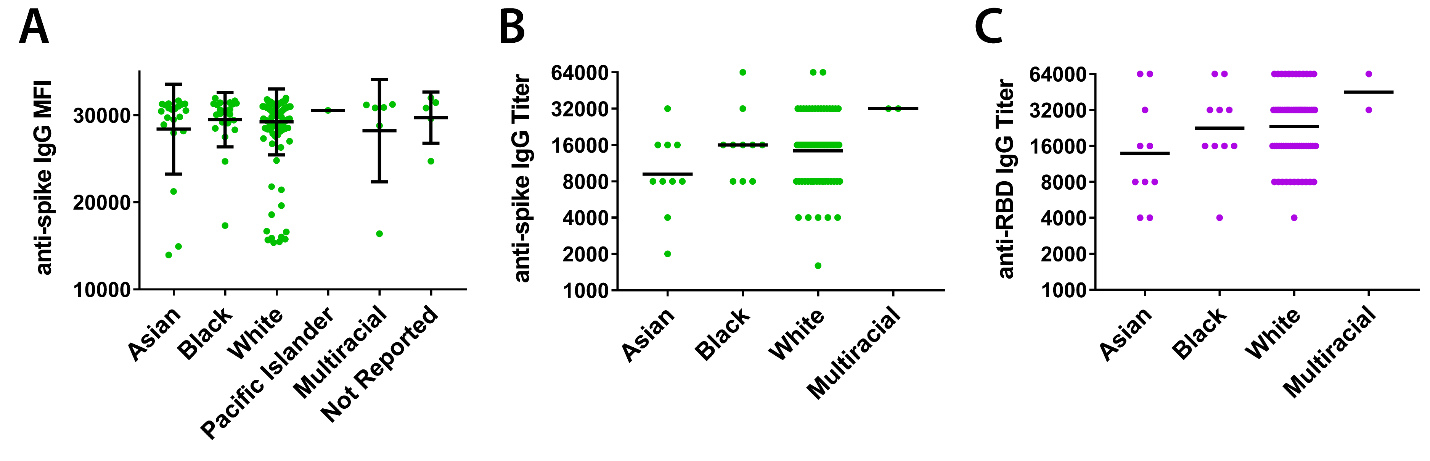

**Supplemental Figure 2: Lack of association between race and BNT162b2-induced anti-SARS-CoV-2 antibody titers.** Samples collected one month (mean 36.8 days) after participants received the second BNT162b2 vaccine dose were tested for antibodies against SARS-CoV-2 spike protein (green) and RBD (purple) and plotted against participant race. (A) Antibody reactivity against SARS-CoV-2 spike protein in samples diluted 1:400, reported as MFI. (B) Endpoint dilution titers of anti-spike IgG and (C) anti-RBD IgG measured in a subset of participants. (ANOVA (A) and Kruskal-Wallis (B,C) analyses were used to assess significance. N = 206 for panel A and n = 101 for panels B and C. Bars indicate mean and standard deviation (A) or geometric mean (B,C). Titers recorded as >32000 are plotted as 64000).

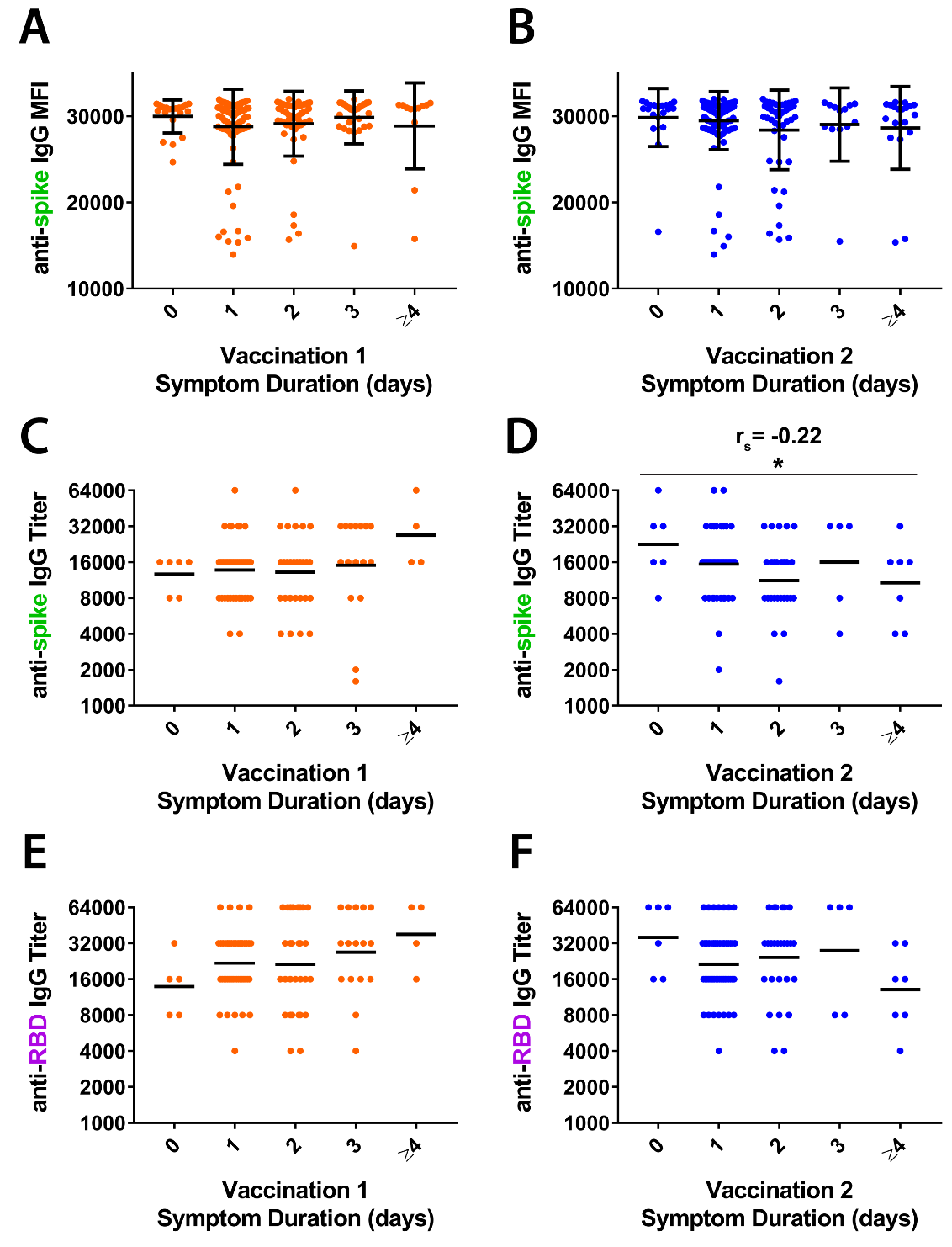

**Supplemental Figure 3: Duration of symptoms and BNT162b2-induced antibody levels.** Samples collected one month (mean 36.8 days) after participants received the second vaccine dose were tested for antibodies against SARS-CoV-2 spike protein and RBD and plotted against total symptom duration. (A, B) Levels of anti-spike IgG antibodies as measured by MFI after first (orange) and second (blue) vaccination (N=206). (C, D) Titers of anti-spike IgG antibodies after first and second vaccination (n=101). (E, F) Titers of anti-RBD IgG antibodies after first and second vaccination (n=101). (Bars indicate mean and standard deviation (A,B) or geometric mean (C-D).Titers recorded as >32000 are plotted as 64000. Spearman rank correlation analysis demonstrated a negative correlation of vaccination 2 symptom duration with anti-spike IgG antibody titers:* p= 0.0274, r_s_ = -0.22).

**Supplemental Methods**

***Antibody testing***

Testing for IgG antibodies against SARS-CoV-2 spike protein and receptor binding domain (RBD) was conducted using microsphere-based multiplex immunoassays (MMIAs) built using Luminex xMAP-based technology as previously described ^14^. Two MMIA antigen-based panels were used, one with trimeric prefusion-stabilized ectodomain spike proteins of SARS-CoV-2 and the four seasonal human coronaviruses (HKU1, OC43, 229E, and NL63), and one with SARS-CoV-2 RBD and nucleocapsid protein. The SARS-CoV-2 spike protein was obtained from LakePharma, Inc (San Carlos, CA, USA), whereas the RBD was provided by Dr. Dominic Esposito (FNLCR, Frederick, MD, USA)^1^ and nucleocapsid protein was obtained from RayBiotech (Peachtree Corners, GA, USA). Sensitivity of this assay for the detection of SARS-CoV-2 IgG antibodies from blood specimens 7-28 days after the onset of symptoms is 94% and specificity is 100% ^2^. Mean fluorescence intensity (MFI) was measured for all serum samples prepared at a dilution of 1:400 using the MMIA panel measuring IgG antibodies against SARS-CoV-2 spike protein. Additionally, endpoint dilution titers were determined for the first 101 participants with one-month post-2^nd^ vaccination serum samples obtained from the study cohort on both MMIA assays. Titers were peformed at seven two-fold dilutions from 1:500 to 1:32000 in phosphate buffered saline (PBS) without calcium and magnesium. Endpoint titers of IgG against SARS-CoV-2 spike and RBD were defined as the last dilution at which the MFI exceeded the threshold cutoff values of 4774 MFI and 4666 MFI respectively, as previously reported in Laing et al ^2^.

**References Supplemental Methods**

1.	Mehalko J, Drew M, Snead K, et al. Improved production of SARS-CoV-2 spike receptor-binding domain (RBD) for serology assays. *Protein Expr Purif*. Mar 2021;179:105802. doi:10.1016/j.pep.2020.105802

2.	Laing ED, Sterling SL, Richard SA, et al. Antigen-based multiplex strategies to discriminate SARS-CoV-2 natural and vaccine induced immunity from seasonal human coronavirus humoral responses. *medRxiv*. Feb 12 2021;doi:10.1101/2021.02.10.21251518
